# Arrayed Imaging Reflectometry monitoring of anti-viral antibody production throughout vaccination and breakthrough Covid-19

**DOI:** 10.1101/2022.11.08.22282042

**Authors:** Alanna M. Klose, Gabrielle Kosoy, Benjamin L. Miller

## Abstract

Immune responses to COVID-19 infection and vaccination are individual and varied. There is a need to understand the timeline of vaccination efficacy against current and yet to be discovered viral mutations. Assessing immunity to SARS-CoV-2 in the context of immunity to other respiratory viruses is also valuable. Here we demonstrate the capability of a fully automated prototype Arrayed Imaging Reflectometry (AIR) system to perform reliable longitudinal serology against a 34-plex respiratory array. The array contains antigens for respiratory syncytial virus, seasonal influenza, common human coronaviruses, MERS, SARS-CoV-1, and SARS-CoV-2. AIR measures a change in reflectivity due to the binding of serum antibodies to the antigens on the array. Samples were collected from convalescent COVID-19 donors and individuals vaccinated with a two-dose mRNA vaccine regimen. Vaccinated samples were collected prior to the first dose, one week after the first dose, one week after the second dose, and monthly thereafter. Information following booster dose and/or breakthrough infection is included for a subset of subjects. Longitudinal samples of vaccinated individuals demonstrate a rise and fall of SARS-CoV-2 spike antibodies in agreement with general knowledge of the adaptive immune response and other studies. Linear Regression analysis was performed to understand the relationship between antibodies binding to different antigens on the array. Our analysis identified strong correlations between closely related influenza virus strains as well as correlations between SARS-CoV-2, SARS-CoV-1, and human coronavirus 229E. A small test of using diluted whole blood from a fingerstick provided clean arrays with antibody binding comparable to serum. Potential applications include assessing immunity in the context of exposure to multiple respiratory viruses, clinical serology, population monitoring to facilitate public health recommendations, and vaccine development against new viruses and virus mutations.

## Introduction

As we are all well aware, the COVID-19 pandemic, caused by the SARS-CoV-2 virus, challenged the social and economic stability of the world since emerging in late 2019 [1]. It motivated the rapid expansion and development of diagnostic methods for identifying infection, including nucleic acid, antigen, and antibody detection [2]. The deployment of safe and effective vaccines in late 2020 changed the course of the pandemic by reducing the severity of disease and suggested the possibility that immunity would be maintained by a future endemic status [3,4]. The virus has subsequently mutated to produce variants of concern (VOCs) that have caused waves of breakthrough infections [5]. A third dose of BNT162b2, the mRNA vaccine developed by Pfizer, has been shown to improve immunity to the omicron variant BA.1 by increasing the neutralizing capability of circulating IgGs [6,7]. Data from a study of a fourth BNT162b2 vaccine dose in Israel suggests that the doubly boosted immune response lasts less than or equal to two months in individuals over 60 years of age [8]. As governments worldwide have begun to embrace an endemic future of COVID-19 it will be important to track immune responses to VOCs in vaccinated individuals to inform decisions about mask wearing, booster doses for healthy and immunocompromised individuals, and development of VOC-specific vaccines [9].

Cross-reactive antibodies against common human coronaviruses (hCoVs), SARS-CoV-1, and MERS are increased after COVID-19 infection [10], and to a lesser extent vaccination [11,12]. However, the role of pre-existing cross-reactive T-cells and antibodies to hCoVs in protection against Covid-19 is controversial, with some studies reporting enhanced immune responses [13,14], and others reporting no protection [15] or even a decreased immune response [16]. The ability to evaluate cross-reactive antibody binding to hCoV antigens, SARS-CoV-1, MERS, and SARS-CoV-2 antigens with a single test could enable further exploration into the relationship between cross-reactivity of the antibodies and disease outcome.

Influenza surveillance around the world currently relies on either RT-PCR or rapid tests. Rapid tests only indicate the presence of Influenza A or B, not subtype or individual strain. RT-PCR can test for subtype and strain but requires primers for each strain that is being tested for [17]. Tracking which strains of influenza are circulating or are closely cross-reactive is important for vaccine development and outbreak prediction [18].

For several years, we have worked to develop array-based methods based on Arrayed Imaging Reflectometry (AIR) for rapidly assessing immunity to upper respiratory viruses [19]. AIR relies on the quantitative perturbation of a near-perfect antireflective condition on a silicon/silicon dioxide chip as targets bind to the arrayed probes [20,21]. As an imaging technique, AIR can quantify binding of more than 100 targets on an array independently and simultaneously [22]. In previous work, we developed an AIR array suitable for monitoring the immune response to SARS-CoV-2, and demonstrated multiplex data consistent with single-analyte ELISA [23]. Here, we have employed a prototype commercial version of AIR, which requires only a few microliters of each serum sample or whole blood, with a significantly expanded array of recombinant proteins (antigens) from upper respiratory viruses. We demonstrate the use of AIR to screen for antibodies against antigens from 34 human respiratory viruses including SARS-CoV-2, SARS-CoV-1, MERS, common hCoVs, and pandemic and seasonal influenza strains of type A and B. This is a longitudinal study of adult subjects who were infected with SARS-CoV-2, received a Pfizer/BioNTech (BNT162b2) or Moderna (MRNA-1273) vaccine, or experienced both infection and vaccination. Vaccinated samples were collected prior to the first dose, one week after the first dose, one week after the second dose. In some individuals we were able to obtain samples monthly for up to 6 months after the second vaccine dose, and two weeks after a breakthrough infection.

This 34-plex array generates 561 combinations of probes, which creates an opportunity to evaluate relationships between antibody responses between probe antigens. Linear regression provides a single numerical measure (coefficient of determination, R^2^ value) of the influence that one variable has on the other. This analysis helps us quantify the impact that the Pfizer and Moderna vaccines, which consist of mRNA encoding the full-length SARS-CoV-2 spike protein [24,25], had on the immune response to other SARS-CoV-2 antigens, including the D614G mutation, as well as related SARS-CoV-1, MERS, and hCoV proteins. This could help identify cross-reactive binding of antibodies to new variants or identify when vaccines need to be modified to improve immune response. Although the arrays tested here are commercially produced, the technology allows for easy modification to add new Covid-19 or influenza VOCs as they arise.

## Materials and Methods

### Human Samples

For serum collection, whole blood was drawn via venipuncture, allowed to clot at ambient temperature for at least 1 hour, and then centrifuged at 1200 x g for 15 min. Serum was drawn off via pipette, aliquoted, and stored at −80 °C prior to use. Sera were drawn under protocols approved by the University of Rochester Medical Center Institutional Review Board for dermatology department assay development. Whole blood assays were performed using pressure activated safety lancets to prick the subject’s finger and pipette 3 μL of blood. Blood was immediately diluted and used in the assay as described below. Sample IDs used in this manuscript were not known to any personnel outside the research group.

### ZIVA by Adarza Biosystems, Inc

ZIVA is an automated version of AIR produced in prototype form by Adarza Biosystems, formerly St. Louis, MO. While Adarza has ceased operations, the methods and results described here are consistent with and transferrable to other AIR instrumentation developed in our laboratory. The ZIVA platform consists of 96-well plates of individually packaged pre-arrayed AIR chips in custom-designed cartridges for sample addition and a fully automated instrument for processing the cartridges through washing, imaging, and data processing (Fig 1). Cartridges came in an Acute Respiratory Virus Array (ARVA) kit also purchased from Adarza Biosystems. Each cartridge accepts 45 μL of sample, which is easily applied using standard multichannel pipettes. In this study, all serum samples were diluted 1:20 in Assay Wash Buffer (AWB: mPBS with 0.005% tween-20, pH 7.2) containing 20% Fetal Bovine Serum (FBS), which means that 3 μL of serum is required for the assay. The samples were added to the cartridges and allowed to incubate for 1 hour at RT, shaking at 420 RPM, plus 2 hours at RT without shaking. The 2 extra hours are not necessary, but were added due to technical difficulties with the prototype instrument during the first group of samples, and were retained for consistency throughout the study. The plate was then loaded into the instrument and processed.

**Fig 1.**
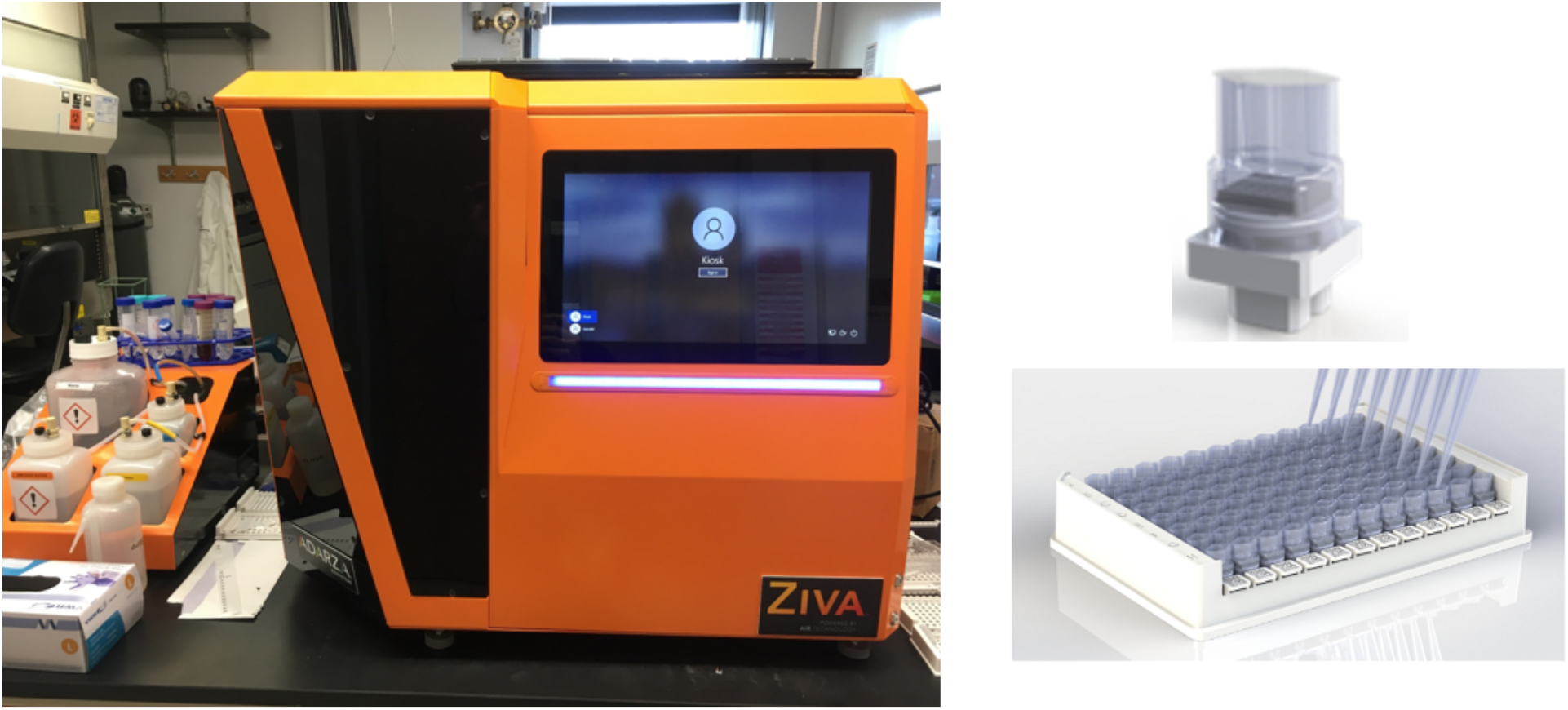
ZIVA system including 96-well plate full of AIR chip cartridges and fully automated instrument.

### Data Processing

AIR provides reflectivity data in an image, where the intensity of each pixel may be converted to thickness via well-established algorithms [21]. Thickness in turn may be converted to concentration if desired with reference to a calibration curve. While the ZIVA instrument has its own software that provides both numerical and graphical results, it also has an option to provide raw reflectivity and converted thickness data in a csv file. The data shown here are the converted thicknesses from that raw csv file which were extracted and organized using custom Matlab [26] and R [27]/R Studio [28] scripts. Heatmaps report thickness change in Ångstroms (Å), and were generated using Pandas [29] and Seaborn [30]. Data preprocessing and plotting were done using tidyverse [31]. Linear regression analysis was done using the stats package and ggpmisc [32]. The thickness change is calculated by subtracting the thickness of each probe on a negative control chip, also referred to as a blank (exposed only to AWB with 20% FBS) from each probe on a sample chip. The limit of detection (LOD) was calculated for each antigen using the raw thickness data from 4 negative control chips (n=4) run on the array when the kit was first opened. The limit of detection was calculated at the mean thickness of the blank chips (μblank) plus 3 times the standard deviation of the blank chips (SDblank).

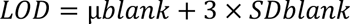

Only those thickness changes above the LOD for each antigen were applied toward linear regression analysis of relationships between antigen responses on the array. Furthermore, any linear regression model with a p-value <0.01 was considered insignificant.

The instrument does not require processing a full plate of samples at a time, and therefore partial plates were used throughout the study in an attempt to preserve available cartridges. This also meant that some samples were processed 6 months after the kit was opened, which resulted in some loss of signal on the array. In these cases, linear regression analysis between samples run on “initial” and “decayed” cartridges was used to generate an equation used to calculate the adjusted thickness changes (Fig S1). In the interest of transparency, the unadjusted thickness changes of all vaccination samples are included in Fig S2.

## Results and Discussion

An early 16-plex version of the ARVA was used for initial experiments (Table 1). The samples beginning with ‘SN’ were drawn from subjects who had had an unknown respiratory illness at some time during late 2019 and early 2020. This was early in the pandemic when tests were in short supply, and only SN028 had a PCR-confirmed case of Covid-19. The ‘HD’ samples were acquired at least 14 days after illness from convalescent COVID-19 patients via the University of Rochester Medical Center’s Healthy Donor protocol.

**Table 1.**
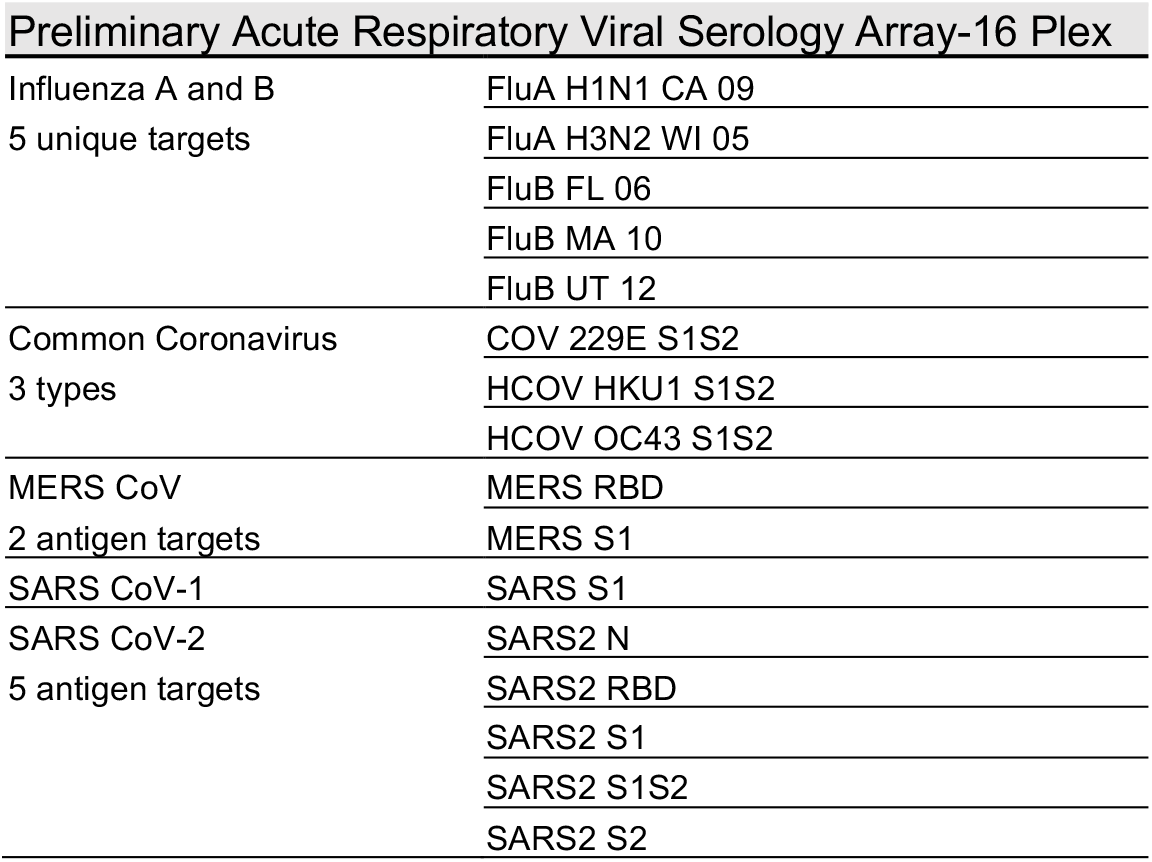
Antigens on an early 16-plex ARVA.

| Preliminary Acute Respiratory Viral Serology Array-16 Plex |  |
| --- | --- |
| Influenza A and B<br>5 unique targets | FluA H1N1 CA 09 |
|  | FluA H3N2 WI 05 |
|  | FluB FL 06 |
|  | FluB MA 10 |
|  | FluB UT 12 |
| Common Coronavirus<br>3 types | COV 229E S1S2 |
|  | HCOV HKU1 S1S2 |
|  | HCOV OC43 S1S2 |
| MERS CoV<br>2 antigen targets | MERS RBD |
|  | MERS S1 |
| SARS CoV-1 | SARS S1 |
| SARS CoV-2<br>5 antigen targets | SARS2 N |
|  | SARS2 RBD |
|  | SARS2 S1 |
|  | SARS2 S1S2 |
|  | SARS2 S2 |

The heatmap in Fig 2 shows antibody binding onto each antigen in the array as a thickness change relative to the control chip (build). We can see varied responses to influenza and common hCoV antigens across individuals, which is expected due to personal health histories and propensity to receive annual flu shots. When looking at the SARS-CoV-2 antigens we see that many, but not all the convalescent patients have antibodies in their serum that bind to N-protein and spike proteins including the receptor binding domain (RBD) as well as full length S1+S2 and individual subunits S1 and S2. This is consistent with work performed on these samples in our laboratory with the preliminary, non-automated version of a SARS-CoV-2 AIR array [23], and with ELISA results on these samples acquired by an independent laboratory [33]. The low SARS-CoV-2 antibody responses in some convalescent patients can be explained by some patients self-reporting illness without a positive PCR test. Assuming that all “SN” samples were uninfected with SARS-CoV-2 with the exception of SN028 (known PCR positive), these values excluding those from SN028 were averaged together into the uninfected group in Fig S3. All HD patients plus SN028 were grouped into the convalescent group. When comparing average antibody build across all convalescent patients to all uninfected patients, there is significantly more antibody binding onto all SARS-CoV-2 antigens in convalescent serum (unpaired, 2-tailed t-test assuming unequal variance). There is also a significant increase in cross-reactive antibody binding to the SARS-CoV-1 S and OC43 S1S2, but not to MERS S1. The significant increase in cross-reactive antibodies against OC43 after infection with COVID-19 is consistent with the ELISA results from the same convalescent samples [33]. Interestingly, that same group provided evidence that hCoV memory B cells against OC43 were activated in response to infection with SARS-CoV-2.

**Fig 2.**
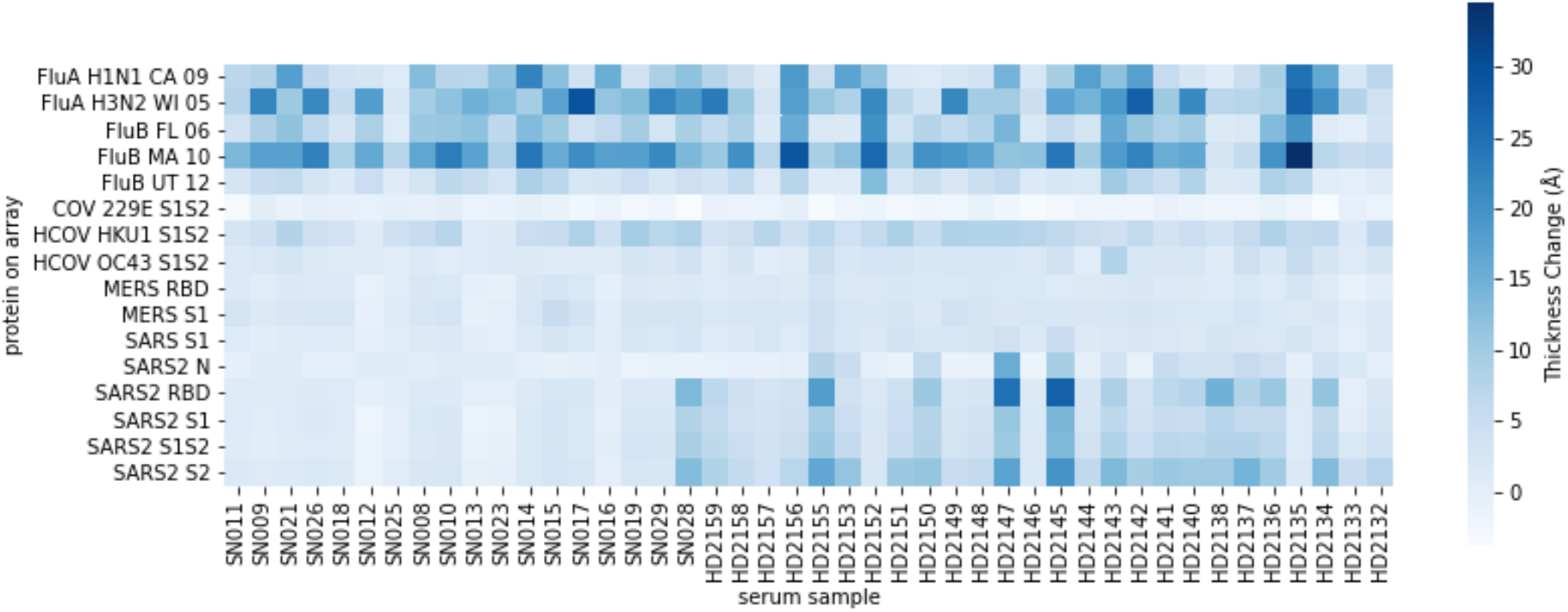
Heatmap of thickness change per array protein. Samples were from convalescent COVID-19 patients (HD) and study subjects (SD) with unknown COVID-19 illness history.

To understand the quantitative range of the assay we selected five high responding convalescent samples and performed serial serum dilutions of 1:20, 1:40, 1:80, 1:160, and 1:320 (Fig 3). Here the data are reported as simply thickness (Å) where negative control information is not subtracted but is instead plotted independently. The standard deviation of serum samples was calculated from replicate probe spots on each array [21]. Positive samples were determined as those significantly different from control by one-tailed two-sample t-test with p < 0.01. The sample dilutions were all analytically well-behaved and titrated to zero. While the creation of a standard curve to convert thickness change to antibody concentration was not within the scope of this study, these data suggest that it would be possible to measure protein concentration using the ZIVA platform.

**Fig 3.**
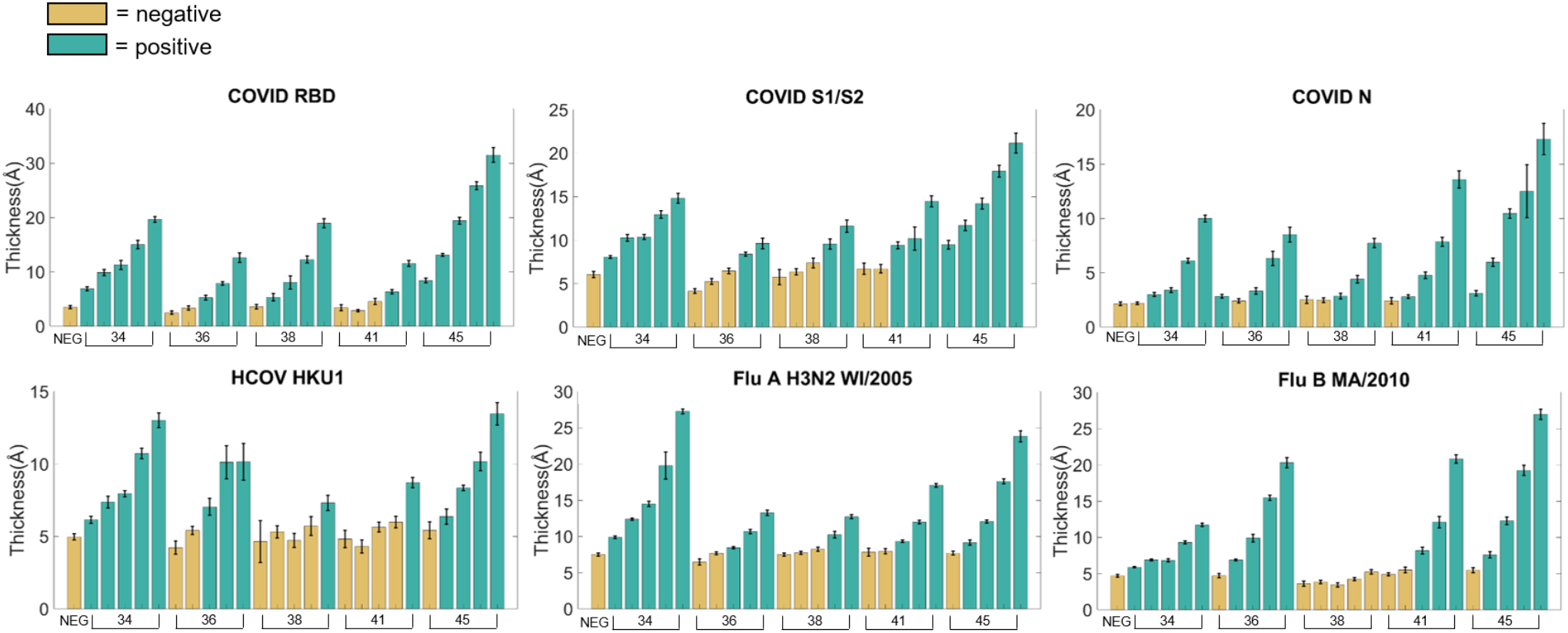
Serial dilutions of selected convalescent serum samples. Dilutions were two-fold (1:20, 1:40, 1:80, 1:160, 1:320) on serum samples HD2134, HD2136, HD2138, HD2141, and HD2145. Positive samples are significantly different from the negative control sample by t-test, p < 0.1.

An expanded 34-plex ARVA kit (Table 2) was used for longitudinal studies of vaccinated individuals (designated VN) in order to track immune response over time. The array generates a large amount of data, making a heatmap the most effective way to gain an overview of the range of responses per sample (Fig 4). The hue indicates thickness change relative to a control chip per antigen on the array per sample, with darker blue indicating a larger thickness change as more antibody bound to antigen.

**Table 2.**
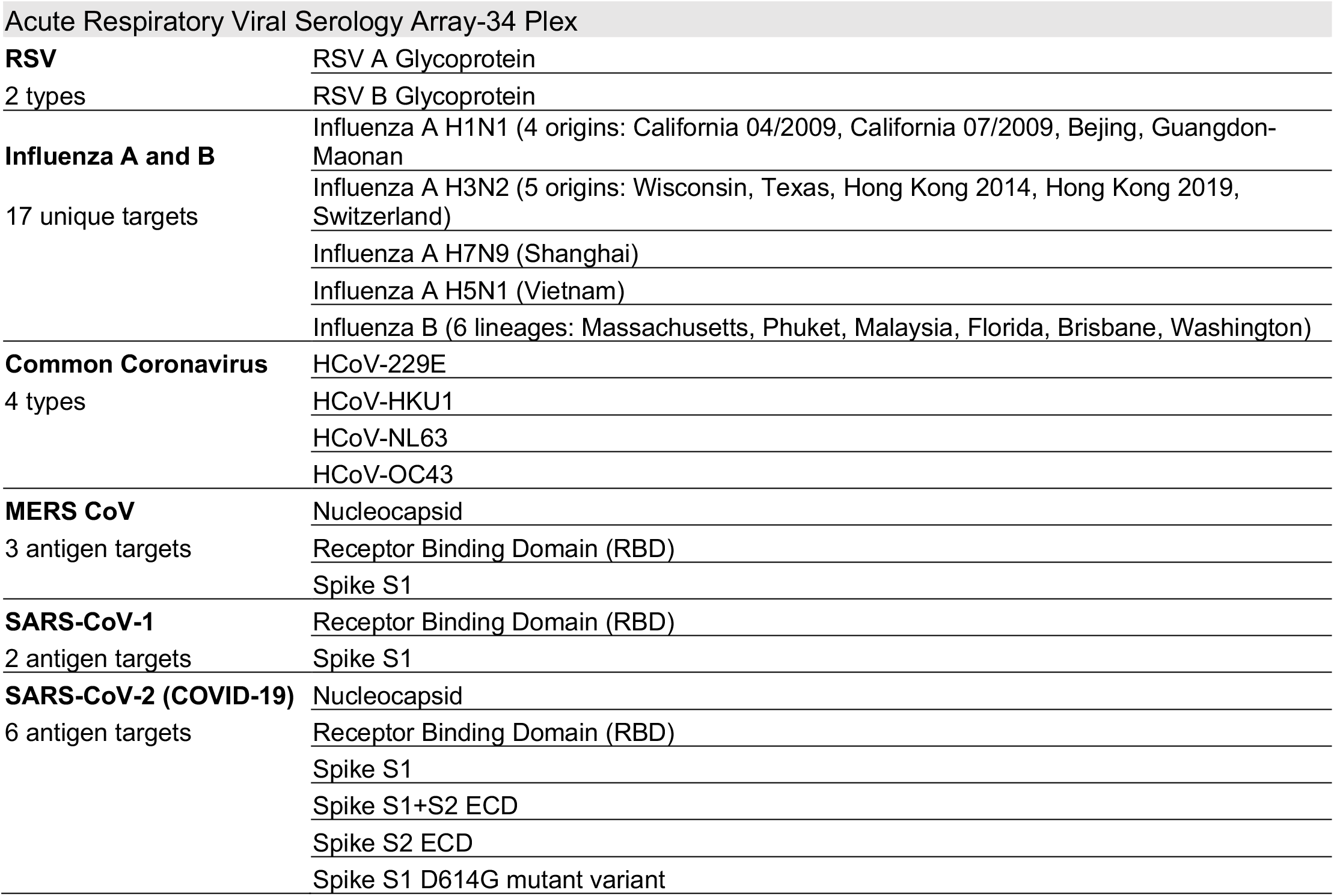
Antigens on a 34-plex ARVA.

|  |  |
| --- | --- |
| Acute Respiratory Viral Serology Array-34 Plex |  |
| RSV | RSV A Glycoprotein |
| 2 types | RSV B Glycoprotein |
| Influenza A and B | Influenza A H1N1 (4 origins: California 04/2009, California 07/2009, Beijing, Guangdong-Maonan |
| 17 unique targets | Influenza A H3N2 (5 origins: Wisconsin, Texas, Hong Kong 2014, Hong Kong 2019, Switzerland) |
|  | Influenza A H7N9 (Shanghai) |
|  | Influenza A H5N1 (Vietnam) |
|  | Influenza B (6 lineages: Massachusetts, Phuket, Malaysia, Florida, Brisbane, Washington) |
| Common Coronavirus | HCoV-229E |
| 4 types | HCoV-HKU1 |
|  | HCoV-NL63 |
|  | HCoV-OC43 |
| MERS CoV | Nucleocapsid |
| 3 antigen targets | Receptor Binding Domain (RBD) |
|  | Spike S1 |
| SARS-CoV-1 | Receptor Binding Domain (RBD) |
| 2 antigen targets | Spike S1 |
| SARS-CoV-2 (COVID-19) | Nucleocapsid |
| 6 antigen targets | Receptor Binding Domain (RBD) |
|  | Spike S1 |
|  | Spike S1+S2 ECD |
|  | Spike S2 ECD |
|  | Spike S1 D614G mutant variant |

**Fig 4.**
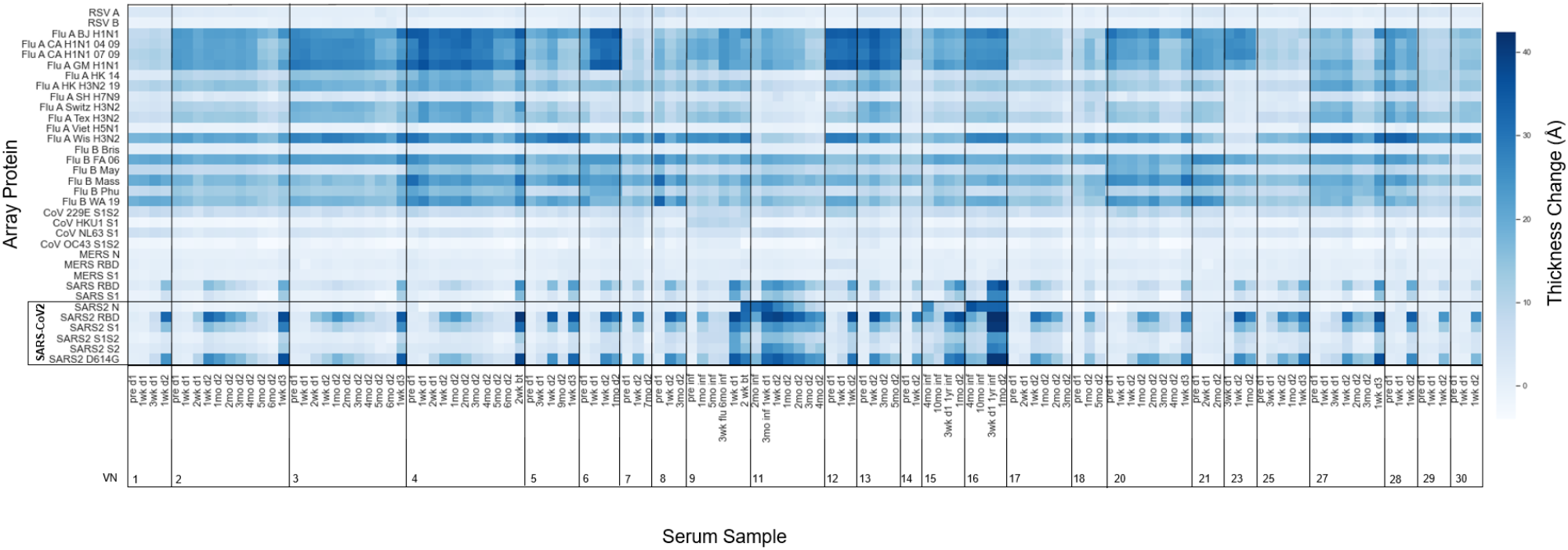
Thickness Change per array protein in vaccinated subjects. Thickness change is representative of antibodies binding to antigens on the 34-plex array for longitudinal serum samples collected from vaccinated subjects.

As expected based on recent studies of time-dependent antibody titers following vaccination [34,35,36,37], the thickness change on the SARS-CoV-2 antigens is dynamic. Looking at VN02, we see that this subject had an increase in SARS-CoV-2 spike antibodies two weeks after the first vaccine dose, which is consistent with other vaccine studies [38]. This pattern holds for other subjects (VN-01, 03, 04, 05, 17, 25, 27) who also donated serum two weeks after the first dose. Additionally, these observations fit with general knowledge of the adaptive immune response. Our results show that the multiplex ARVA array is able to differentiate subjects who had a covid-19 infection (9, 11, 15, and 16) from those who only received the vaccine, by looking at the thickness change caused by antibodies binding to the N-protein. The N-protein is located inside of the viral envelope and interacts with the viral RNA [39]. Since the vaccine mRNA only encodes the spike protein, there isn’t an immune response to N-protein in naïve vaccinated individuals. A few of these subjects (VN-21, 23, and 30) were taking immunosuppressive medications during the course of the study. While VN23 and VN30 seem to mount immune responses to the vaccine, the same cannot be said of VN21. This is a very small sample size, but suggests that AIR technology and the ARVA array could be used to screen immunosuppressed patients to determine if they need extra vaccine doses, antibody therapy, or if they need to take extra precaution to avoid infection. Likewise, such screening could bring peace of mind to immunosuppressed patients who do generate antibodies in response to vaccination, especially since antibody titer correlates with neutralizing capability and protection against severe disease [40].

Antibody response due to vaccination was assessed by grouping samples acquired before vaccination and samples acquired 1-2 weeks after the second vaccine dose (Fig S4). There are significant increases in antibodies binding all SARS-CoV-2 antigens except the N-protein as expected, since the vaccines encode the full-length spike protein, and significant cross-reactive antibodies to SARS-CoV-1 RBD and S1, MERS-CoV-S1, and 229E. Recent work profiling cross-reactive antibodies to related coronaviruses after vaccination of naïve individuals found significant antibody cross-reactivity to SARS-CoV-1 and MERS-CoV spike proteins [12]. Cross-reactivity of antibodies toward common cold hCoVs following vaccination against SARS-CoV-2 has also been observed [11]. It seems that most of this cross-reactivity is recognizing conserved epitopes on the S2 subunit of these hCoVs [10,11]. Our array only included the S1 subunit of NL63 and HKU1. This array included the full-length spike protein for OC43, but many of the responses were below the limit of detection for that protein on this version of the array.

The build onto the influenza antigens is fairly consistent for many subjects, while others have increases in antibody binding. VN09 received a flu shot prior to the fourth serum sample as noted in the figure. While the 2020-2021 quadrivalent flu shot was designed to activate an immune response against A/Guangdong-Maonan/SWL1536/2019 (H1N1) pdm09-like virus, A A/Hong Kong/2671/2019 (H3N2)-like virus, B/Washington/02/2019-like virus (B/Victoria lineage), and B/Phuket/3073/2013-like virus (B/Yamagata lineage) [41], this individual seemed to mount an immune response to other influenza antigens on the array, which is probably cross-reactivity among influenza strains. This may be the case for other subjects as well, but we lack information about flu shots or illness history to be able to confirm.

Even though information about influenza vaccination or infection during the time frame that these samples were collected is not available, there is still an opportunity to look for correlated immune responses to antigens across the array. Linear regression analysis provides an R^2^ value indicating how much influence one variable has on the other. In this context we are using it to determine the correlation between samples, where a perfect correlation has R^2^ equal to 1. Biologically, stronger correlations mean that higher amounts of antibodies binding to the epitope of one protein indicate more antibodies binding to the epitope of the second protein. Linear regression was only performed on thickness change data that was larger than the calculated limit of detection (LOD) for each protein on the array, and only the models with p-values < 0.01 were considered representative of a true relationship between variables. Of note, there were no negative correlations between any of the antigens on the array, indicating that vaccination with these SARS-CoV-2 vaccines doesn’t negatively affect the immune response to other upper respiratory viruses. As a positive control, we saw that the two pandemic strains of influenza isolated in California in the same year (Cal09) had a correlation of 1 (Fig 5). This is expected because these two proteins have a nucleic acid sequence similarity of 99.9%. We were able to identify how strongly the subtypes, H1N1, H3N2, and influenza B, influenced each other. Proteins of the same subtype showed very strong correlation, with Influenza A Beijing H1N1 and Influenza A California H1N1 07-2009 showing the strongest relationship (R^2^ = 0.96). Influenza A Texas H3N2 and Influenza A Hong Kong H3N2 2019 were the most strongly correlated H3N2 subtypes (R^2^=0.83). Influenza B Washington 2019 and Influenza B Massachusetts were the most strongly correlated influenza B strains with R^2^=0.83.

**Fig 5.**
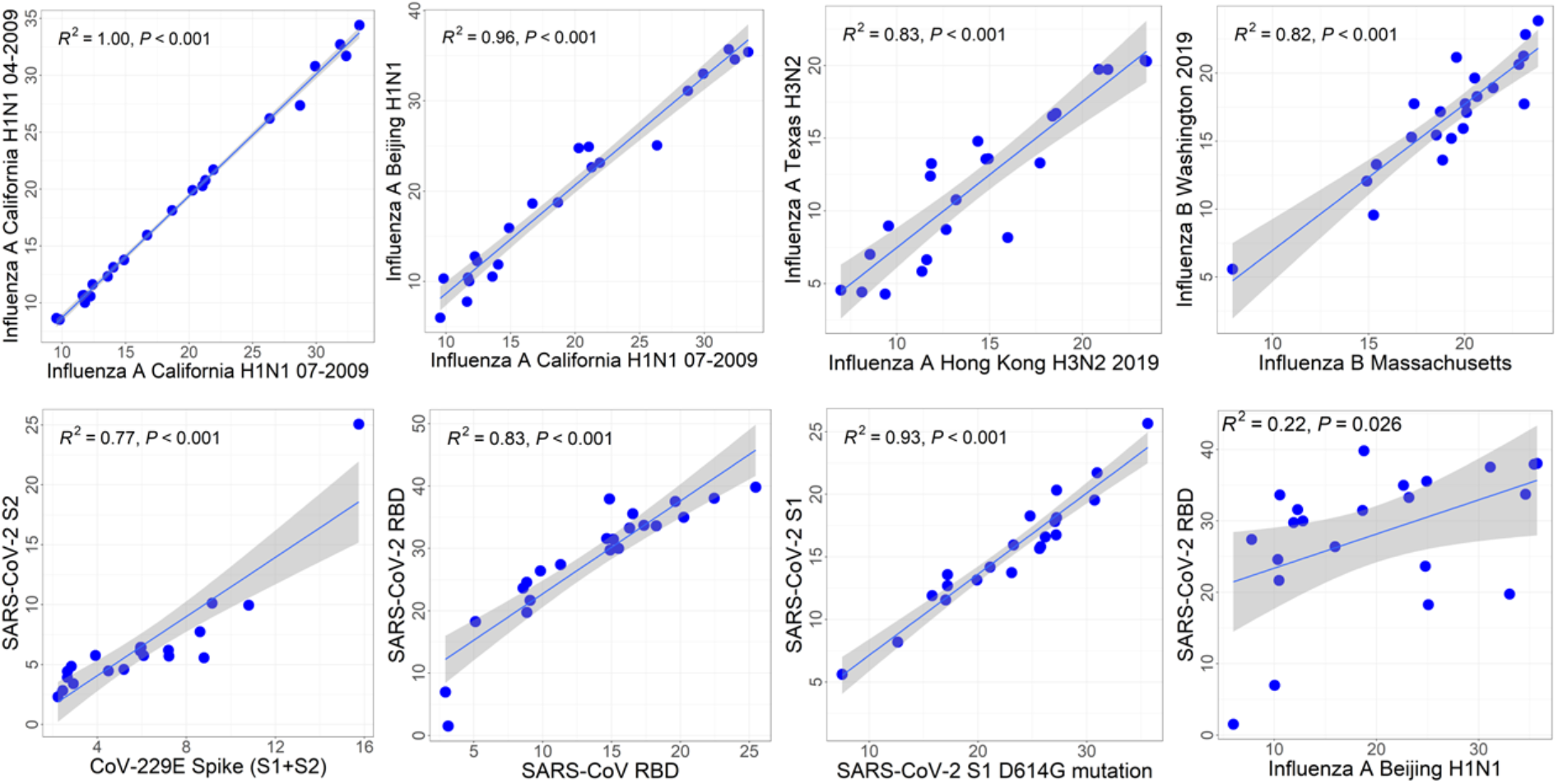
Selected correlations between proteins by linear regression. Plots were made with data from samples drawn within two weeks of the second dose of a SARS-CoV-2 mRNA vaccine. Points are individual samples, and lines are linear regressions with a grey band indicating standard deviation.

There is a strong correlation between antibody binding to the 229E full length spike protein and SARS2-S2 subunit (R^2^=0.77) on our array. The alpha-coronavirus 229E has 31% amino acid sequence similarity for the spike protein of beta-coronaviruses SARS-CoV-2 and SARS-CoV-1, particularly in the conserved regions of the spike protein [42]. Titers of 229E antibodies have been shown to increase in convalescent and immunized populations, and antibody binding to 229E decreased 71% after depleting the serum with a monomeric SARS-CoV-2 S2 subdomain [11,33].

There is also a strong correlation between SARS-CoV-1 RBD and SARS-CoV-2 RBD (R^2^=0.83). This is expected because these peptides share 74% amino acid sequence identity [43]. The D614G spike mutation became widespread after March 2020 [44]. Studies of the structure of the S1 and D614G S1 variant indicate that the D614G variant could have more antibody binding due to having a more flexible S1-S2 interface [45]. Antibody neutralization studies have shown slightly more antibody neutralization against the D614G variant [46]. On our array, D614G S1 protein is observed to have more binding overall than the wild-type S1 (Fig S4), and the correlation between them is very strong (R^2^=0.93) (Fig 5). Many antibody-binding responses are the array were not strongly correlated. For example, SARS-CoV-2 RBD and Influenza A Beijing hemagglutinin do not have a strong relationship, which is expected because these viruses are phylogenetically distinct.

Comparing antibody responses across all of the proteins on our array from longitudinal samples enables comparisons of antibody duration and waning. Fig 6 presents SARS-CoV-2 RBD and influenza strain A/California/07/2009 hemagglutinin protein antibody levels over time. The individuals included in this analysis met the criteria of having a baseline sample collected immediately before receiving the vaccine and samples collected one week after first dose and one week after second dose of vaccine. Some individuals also went on to have monthly blood draws thereafter. Most of these individuals had not been infected with Covid-19 prior to this study, with the exception of VN11. Circulating SARS-CoV-2 RBD antibody levels in VN11 were initially higher than the other, naïve subjects shown in this figure, and immediately increased one week after receiving the first vaccine dose. The naïve subjects did not show an increase in circulating antibodies against SARS-CoV-2 RBD one week after the first vaccine dose, but had a robust response after the second dose, which is consistent with findings from clinical trials [34,35]. The waning of SARS-CoV-2 RBD antibodies over time agrees with other studies showing circulating antibody levels returning close to baseline after 6 months or around 200 days [47]. At day 0, the variance in the number of antibodies present against influenza is greater than against SARS-CoV-2 RBD, but the overall level of influenza antibodies was significantly higher. This was expected because these strains of influenza infections and vaccines have been circulating for over a century, and most if not all individuals have been exposed to them throughout their lifetimes. In contrast, SARS-CoV-2 is a new strain of coronavirus. People who receive influenza vaccinations generally retain influenza-specific IgG antibodies for around two years [48], and cross-reactive antibodies to other strains will make this appear higher.

**Fig 6.**
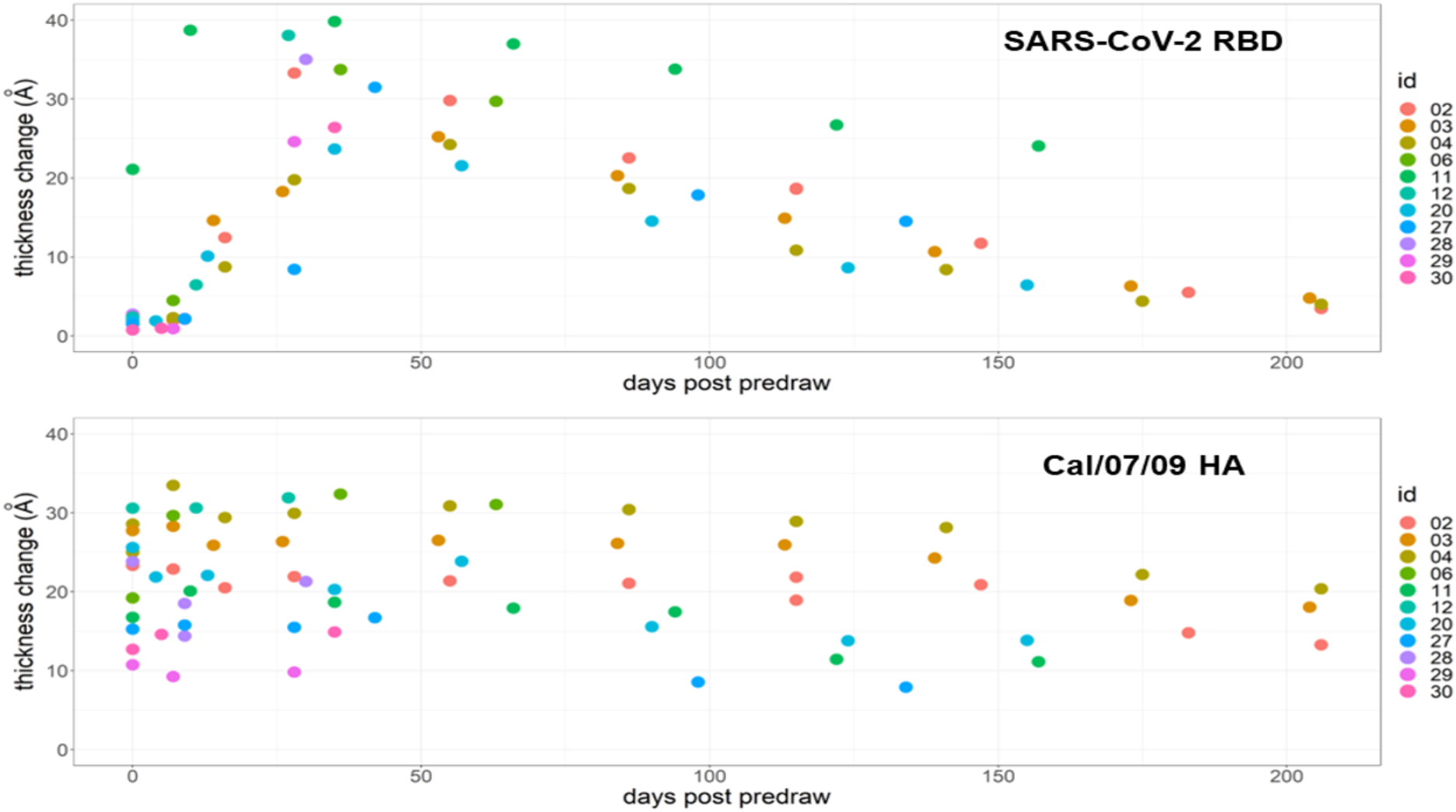
Antibody duration and waning following COVID-19 vaccination. The eleven subjects plotted here (coded by color) all followed the criteria of having a baseline sample collected before receiving the vaccine and samples collected one week after first dose and one week after second dose of vaccine. Day 0 is the baseline value. Circulating antibodies against the SARS-CoV-2 RBD protein increased with vaccination and decayed over time. Antibody response against the influenza strain A/California/07/2009 hemagglutinin protein had larger variance between samples and response remained similar over time.

A subset of subjects who received a booster vaccine dose or had a breakthrough infection is shown in Fig 7. Circulating antibody levels increase from first to second dose, followed by a slow decline over the following six months, and a sharp increase after booster or breakthrough infection. Subjects VN04 and VN09 both had breakthrough infections. Although we expected to see antibodies against the N-protein after infection in both samples, this was not the case for VN04. A low humoral response of antibodies against the N-protein in a fraction of individuals after infection has been observed in other studies [49,50], and younger people with asymptomatic or mild cases tend to produce lower antibody titers against the N-protein [51]. Subject VN09 (a female, under 40, with a mild case of COVID-19) is intriguing because she had a mild PCR-confirmed case of COVID-19 before receiving the vaccine, but didn’t have a robust antibody response to the N-protein until after breakthrough infection. Her second infection seemed to boost her initially low immune response towards the N-protein.

**Fig 7.**
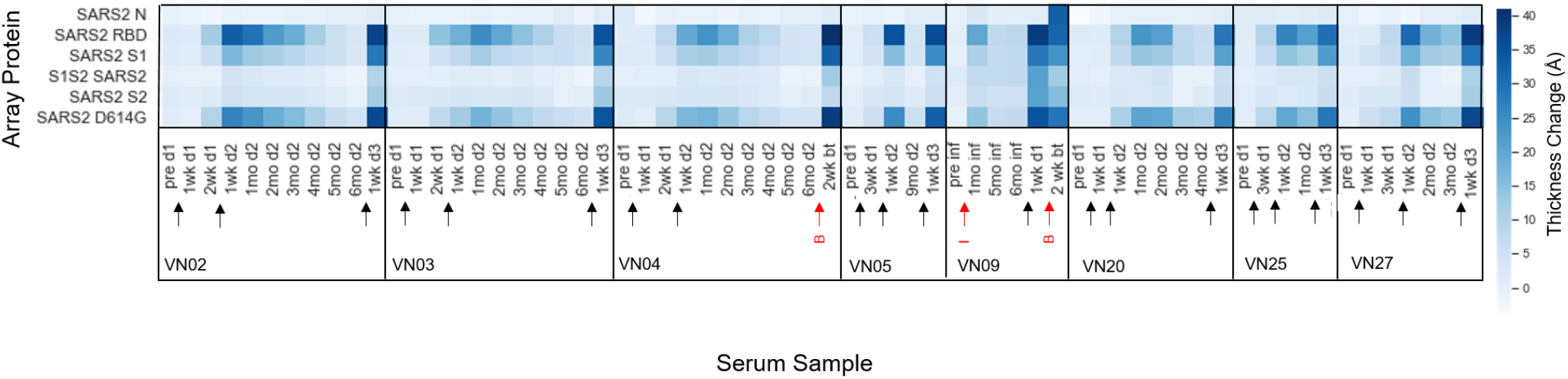
Heatmap of thickness change for SARS-CoV-2 proteins in COVID-19 vaccinated subject serum. Subjects received a booster shot or had a breakthrough infection. Black arrows indicate vaccine doses. Red arrows indicate infection (“I” for infection prior to vaccine, and “B” for breakthrough infection).

The small volume requirements of the ZIVA AIR system make it possible to do a whole blood assay using only a fingerstick volume of blood (3 μL). An initial test produced clean arrays that were comparable to serum in both background and target reflectivity (Fig 8A). The serum and the blood were from the same individual, who had a breakthrough case of COVID-19 confirmed by a positive commercial antigen test. The thickness change on cytC (negative control antigen) and FluA/HK/H3N2/19 was identical between the serum and whole blood. The serum was collected 5 weeks prior to the whole blood sample, so the decrease in circulating antibodies against SARS-CoV-2in the whole blood sample compared to serum is consistent with that seen in figures 4,6, and 7 (Fig 8B).

**Fig 8.**
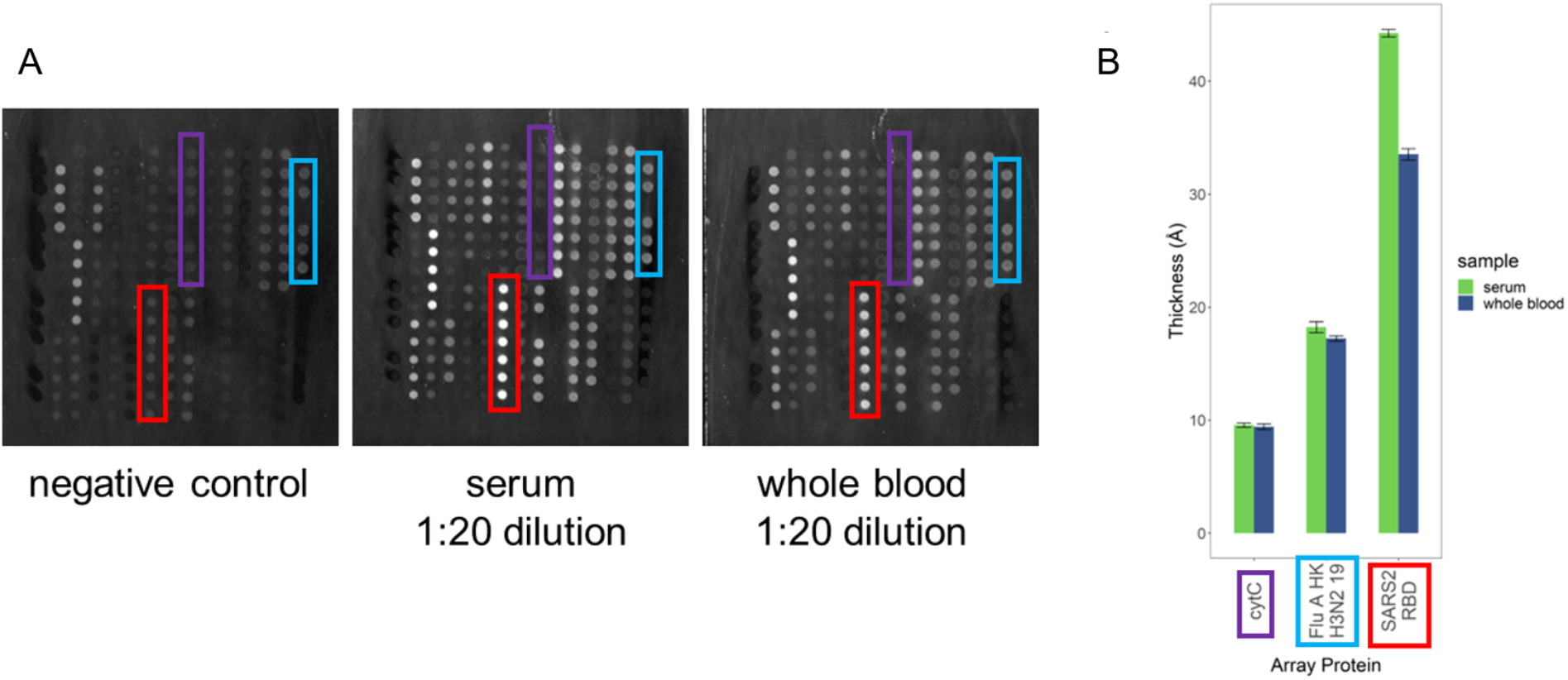
Whole blood assay. (A) Arrays incubated with serum vs. 3 μL whole blood from the same individual. Serum sample was drawn 5 weeks prior to whole blood sample. (B) Thickness increase for three antigens on the array. Error bars are standard deviation of replicate spots of each protein on an array.

## Conclusion

In this study, we have demonstrated that a prototype automated version of the label-free AIR sensor technology is able to profile human antibody responses to 34 antigens from upper respiratory viruses including SARS-CoV-2 from a small (<10 μL) sample of serum or whole blood. Of particular current interest, this approach proved useful in providing insight into immune responses following Covid-19 infection and vaccination. The results were consistent with previous low-multiplex work performed by our lab and with findings elsewhere. Given that AIR is expandable to include 100 or more probes, we can envision future work in which antigens from new SARS-CoV-2 variants are added to the array as they arise. For example, we know now that the booster dose can improve antibody neutralization of the Omicron variant [6,7], but it is not currently known how well existing immunity will adapt to future variants. We expect our tool will be useful to help predict vaccinated immune responses to newly discovered VOCs before they become widespread. Multiplex AIR technology could also be useful for influenza and coronavirus surveillance and could ease further investigation into the relationship between antibody cross-reactivity and disease outcome. Finally, the ability to use a fingerstick quantity of blood to generate a real-time profile of circulating antibodies could be useful as a clinical diagnostic technique. Studies along these lines are currently in progress in our laboratory.

## Data Availability

All data produced in the present study are available upon reasonable request to the authors.

## Acknowledgement

This research was supported by the New York State Empire State Development Fund, and by the U.S. Department of Defense under AIM Photonics, Air Force Contract FA8650-15-2-5220. The views and opinions expressed in this paper are those of the authors and do not reflect the official policy or position of the United States Air Force, Department of Defense, or the U.S. Government. We thank Professor Matthew Brewer, Professor Mark Sangster, and Micah Wiesner for helpful discussions in immunology, virology, data analysis, and programming

