## Supplementary Information for "Arrayed Imaging Reflectometry monitoring of anti-viral antibody production throughout vaccination and breakthrough Covid-19"

### Supporting Information

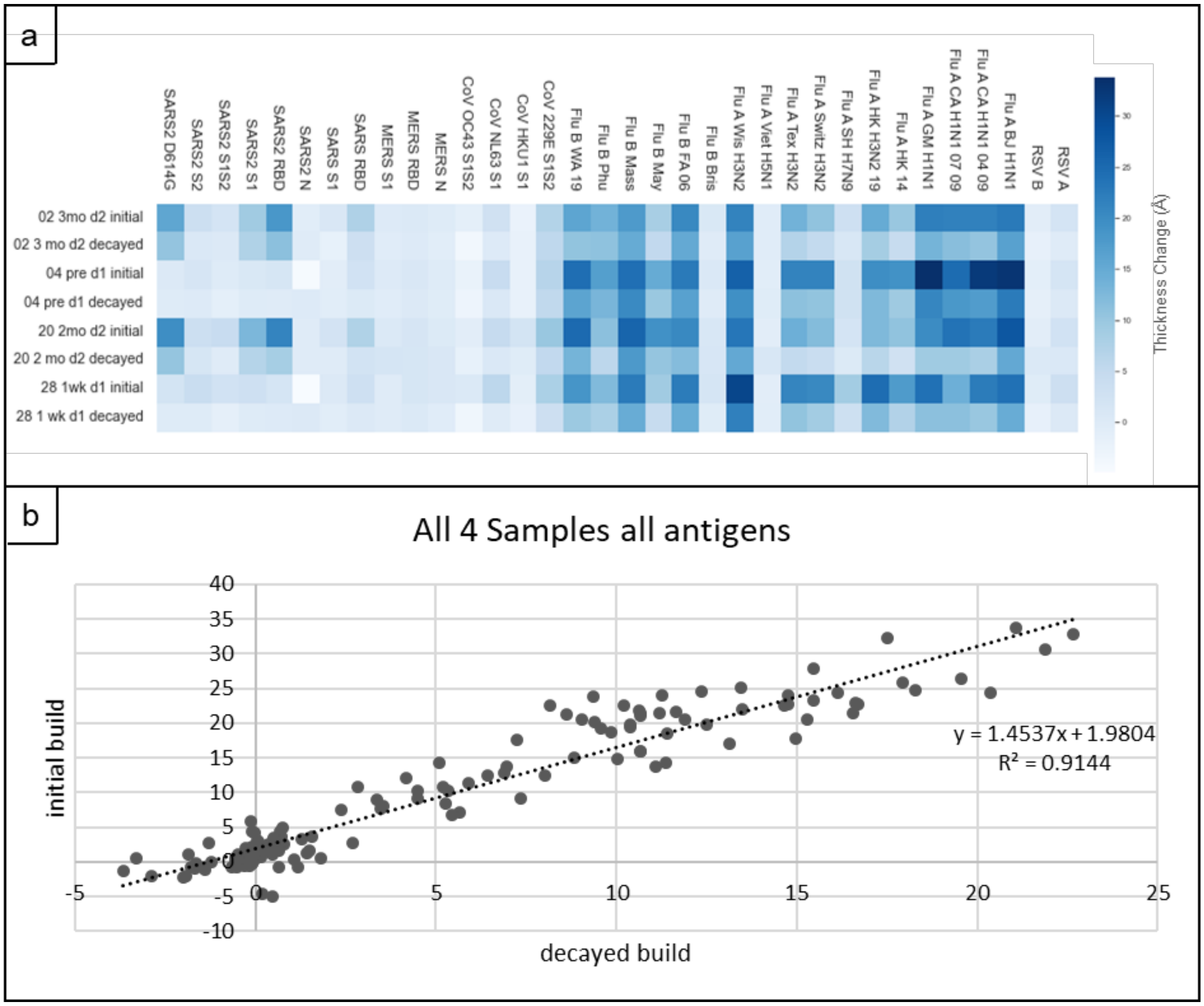


Figure S1. Comparison of the four samples run on both the newly opened 34-plex ARVA array (initial) and the same array after 6 months of the array stored in the 4°C refrigerator (decayed). The samples were different aliquots stored in the -80°C freezer. The thickness change is clearly muted on the decayed array (a). All antigen thickness changes (build) for all samples were plotted to find a relationship between build on the initial array and build on the decayed array. Linear regression demonstrated a strong relationship ($R^2$=0.91), and the equation was used to adjust all of the builds run on the 6-month-old array back up to initial levels (b).

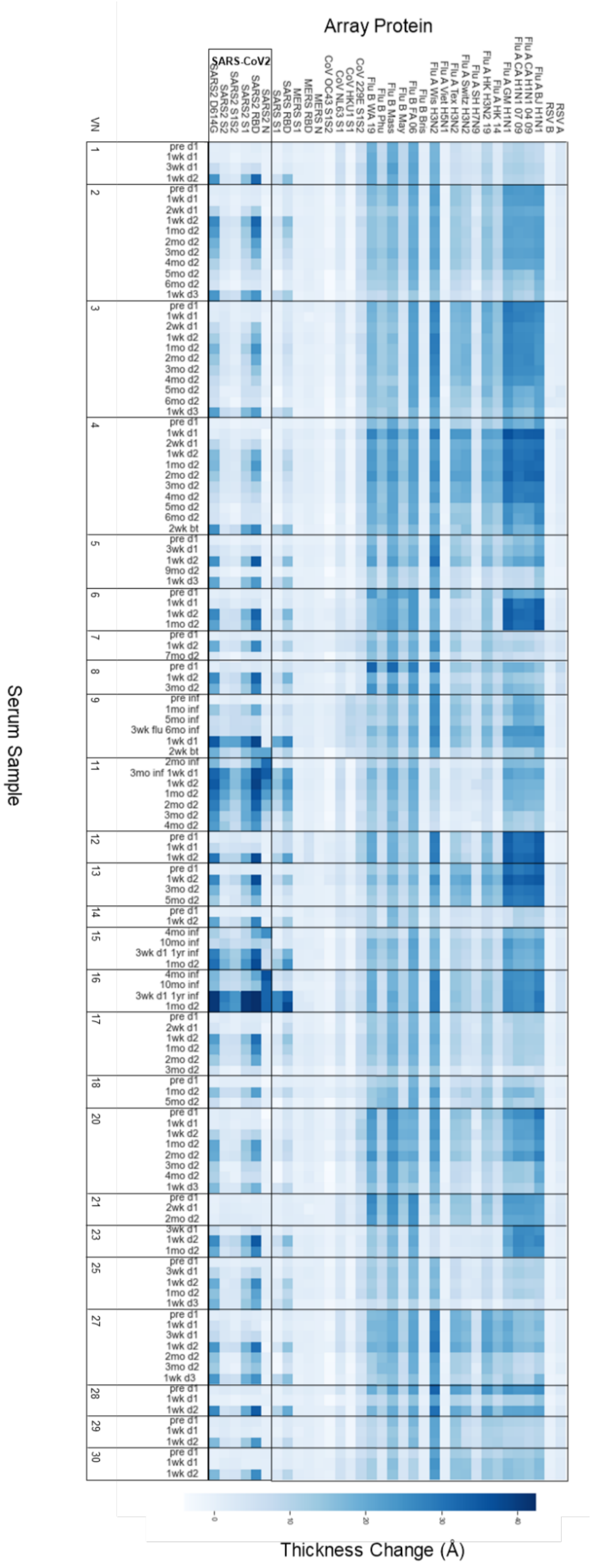


Figure S2. All unadjusted thickness changes for the longitudinal vaccine samples shown in figure 4 of the main text.

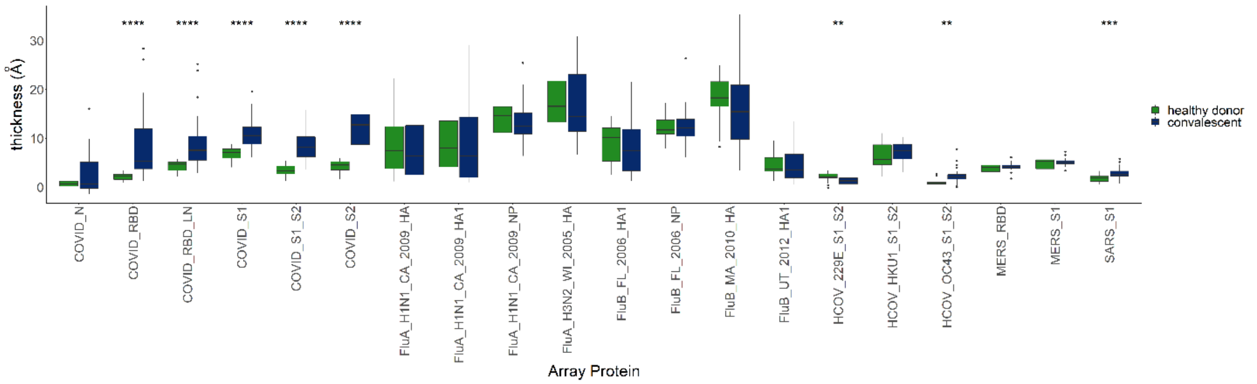


Figure S3. Box plots of thickness (Å) due to antibody binding in Covid-19 convalescent serum vs serum from individuals uninfected with Covid-19 on the 16-plex ARVA array. Boxplots display the median value and data points between the 25th and 75th percentile (boxed), the minimum and maximum data points (tails) along with potential outliers (dots). Significance determined by unpaired, two tailed t-test assuming unequal variance, ****p<0.0001***p<0.001, **p<0.01, *p<0.05.

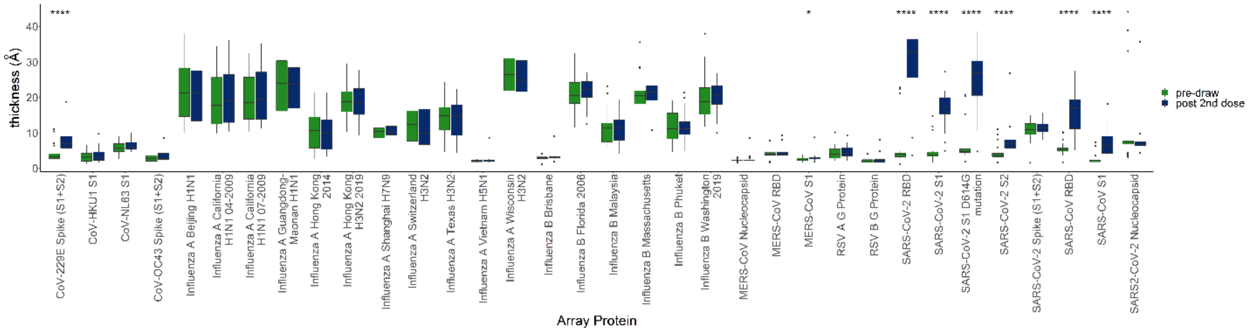


Figure S4. Box plots of thickness (Å) due to antibody binding after incubation in serum from individuals prior to vaccination (no previous infection with Covid-19) and 1-2 weeks after second dose of either a Pfizer/BioNTech (BNT162b2) or Moderna (MRNA-1273) vaccine. Boxplots display the median value and data points between the 25th and 75th percentile (boxed), the minimum and maximum data points (tails) along with potential outliers (dots). Significance determined by paired, two tailed t-test assuming unequal variance, ****p<0.0001***p<0.001, **p<0.01, *p<0.05.

| Influenza strain | Full length name |
|---|---|
| Influenza A Beijing H1N1 | A/Beijing/22808/2009 |
| Influenza A California H1N1 04-2009 | A/California/04/2009 |
| Influenza A California H1N1 07-2009 | A/California/07/2009 |
| Influenza A Guangdong-Maonan H1N1 | A/Guangdong-Maonan/SWL1536/2019 |
| Influenza A Hong Kong 2014 | A/Hong Kong/4801/2014 |
| Influenza A Hong Kong H3N2 2019 | A/Hong Kong/2671/2019 |
| Influenza A Shanghai H7N9 | A/Shanghai/1/2013 |
| Influenza A Switzerland H3N2 | A/Switzerland/9715293/2013 |
| Influenza A Texas H3N2 | A/Texas/50/2012 |
| Influenza A Vietnam H5N1 | A/VietNam/1203/2004 |
| Influenza A Wisconsin H3N2 | A/Wisconsin/67/2005 |
| Influenza B Brisbane | B/Brisbane/60/2008 |
| Influenza B Florida 2006 | B/Florida/4/2006 |
| Influenza B Malaysia | B/Malaysia/2506/2004 |
| Influenza B Massachusetts | B/Massachusetts/03/2010 |
| Influenza B Phuket | B/PHUKET/3073/2013 |
| Influenza B Washington 2019 | B/Washington/02/2019 |

Table S1. Full length names of Influenza strains used in the Ziva arrays.
